# Validation of a treatment selection algorithm for optimal choice of SGLT2 and DPP4 inhibitor therapies in people with type 2 diabetes across major UK ethnicity groups

**DOI:** 10.1101/2025.08.21.25334148

**Authors:** Laura M Güdemann, Katherine G Young, Pedro Cardoso, Bilal A Mateen, Rury R Holman, Naveed Sattar, Ewan R Pearson, Andrew T Hattersley, Angus G Jones, Beverley M Shields, John M Dennis, the MASTERMIND consortium

**Affiliations:** Clinical and Biomedical Sciences, University of Exeter Medical School, Exeter, UK; School of Life Sciences, University of Birmingham, Birmingham, UK; Diabetes Trials Unit, Radcliffe Department of Medicine, University of Oxford, Oxford, UK; NIHR Oxford Biomedical Research Centre, Churchill Hospital, Oxford, UK; Institute of Cardiovascular and Medical Sciences, University of Glasgow, Glasgow, UK; Division of Diabetes, Endocrinology and Reproductive Medicine, Ninewells Hospital and Medical School, University of Dundee, Dundee, UK

## Abstract

**Background:** Targeting type 2 diabetes treatments based on the routine clinical features of individual patients represents a practical precision medicine approach with potential worldwide applicability. We aimed to evaluate a recently proposed treatment selection model predicting differences in glycaemic responses to SGLT2-inhibitors and DPP4-inhibitors across major UK ethnicity groups.

**Methods:** We externally validated the SGLT2i-DPP4i model in UK primary care cohort (CPRD Aurum, 2013-2020) independent of the original model development cohort. Non-insulin treated individuals with type 2 diabetes were identified and categorised by major UK ethnicity groups: White, Black, South Asian and Mixed/Other. For each ethnicity group, we applied a closed testing procedure to assess whether model recalibration was required. After model updates, we assessed the calibration accuracy of predicted differences in glycaemic response (6-month change in HbA1c) between SGLT2i and DPP4i for each ethnicity group.

**Results:** SGLT2i (n=57,749) and DPP4i (n=87,807) initiations were identified amongst people of White (n=114,287; 78.5%), Black (n=6,663; 4.6%), South Asian (n=20,969; 14.4%) and Mixed/Other (n=3,637; 2.5%) ethnicities. Minor model adjustment was required to adjust for greater observed than predicted glycaemic responses to DPP4i (White -1.6 mmol/mol; Black -3.0 mmol/mol; South Asian -2.6 mmol/mol; Mixed/Other -2.6 mmol/mol). SGLT2i predictions did not require adjustment for non-White ethnicity groups. After model updates, average predicted HbA1c reduction was 3.7 mmol/mol (95%CI 3.5-3.9) greater with SGLT2i than DPP4i for those of White ethnicity; this was greater than for those of South Asian (2.1 mmol/mol (95%CI 1.6-2.6)), Black (0.6 mmol/mol (95%CI 0.5-1.7)) and Mixed/Other (2.6 mmol/mol (95%CI 1.4-3.8)) ethnicity groups. For all ethnicity groups, calibration of predicted differential glycaemic treatment effects were well-calibrated.

**Conclusion:** A treatment selection model for SGLT2-inhibitor and DPP4-inhibitor therapies is accurate for all major UK ethnicity groups. Simple recalibration is beneficial to optimise performance and this is recommended prior to deployment of the model in new populations and settings.

## Introduction

Individualised treatment recommendations for intensifying glucose-lowering therapy to improve glycaemic control in people with type 2 diabetes (T2D) are currently lacking. [1] There are a number of different treatment options recommended after first-line metformin, but little guidance to aid prescribing decisions for the majority of patients. Treatment decisions are therefore challenging and therapeutic inertia is a major problem. [2] A precision medicine approach, which utilises individual characteristics, can help tailor treatment selection to target those most likely to benefit. This strategy aims to optimise outcomes by personalising care based on specific patient profiles. Dipeptidyl peptidase-4 inhibitor (DPP4i) and Sodium-Glucose Cotransporter-2 inhibitors (SGLT2i) are recommended for treatment intensification after metformin [3] and represent around 60% of treatment initiations with a second-line treatment in the UK. [4] Hence, their direct comparison in prediction models is clinically relevant.

In a recent study, we developed and validated a novel treatment selection algorithm for SGLT2i and DPP4i therapies which represent around 60% of treatment initiations with a second-line treatment in the UK. [4] This treatment selection model provides individualised estimates of relative glucose-lowering effect (6-month HbA1c response) based on five routinely measured clinical features (baseline HbA1c, current age, body-mass-index (BMI), estimated glomerular filtration rate (eGFR), and alanine aminotransferase (ALT)). [5] It was developed in a large UK population-based cohort and validated in head-to-head randomised clinical trials. [5] Validation showed that the proposed treatment selection model can identify a subgroup of around 40% of people of the study cohort who experience a glycaemic benefit of 5 mmol/mol or greater on SGLT2i compared with DPP4i. A smaller group of around 15% of patients with a greater glycaemic benefit and a lower risk of short-term discontinuation with DPP4i.

A limitation of the algorithm was that model performance was not evaluated across people of different ethnicities due to the relatively low numbers of individuals of non-White ethnicities in the original study datasets. [5] It is therefore important to assess the validity of the proposed treatment selection model across different ethnicity groups to lower risk of algorithmic bias and potential suboptimal or inequitable treatment recommendations for minority populations. [6-8] Evidence from previous studies investigating possible treatment effect heterogeneity for different ethnicity groups is sparse but, some studies suggest that, whilst glycaemic reductions with the SGLT2i class may not vary by ethnicity, [7, 9-12] DPP4i glycaemic reductions may be greater in people of Asian ethnicity compared with people of White ethnicity [13] or non-Asian patient subgroups [14]. We, therefore, aimed to perform an external validation study to assess and optimise the performance of the SGLT2i and DPP4i therapy treatment selection algorithm across major UK ethnicity groups.

## Methods

### Research Design and Participants

Routine clinical data from 960 UK primary care practices from January 1^st^, 2013 to November 6^th^ 2020, were accessed from Clinical Practice Research Datalink (CPRD) Aurum. These data comprised a distinct set of individuals to our development set (CPRD Gold), as the two datasets were collected from different GP practice electronic patient record software systems. [15] Study eligible patients were identified using the same inclusion criteria as previously described: individuals with type 2 diabetes, not insulin-treated, eGFR >45, baseline HbA1c >53 and <120 mmol/mol. [5] Further details on cohort construction and eligibility criteria are given in sFigure 1, with the underlying code available at https://github.com/Exeter-Diabetes/CPRD-LauraJohn-SGLT2iDPPi-ethnicity.

Ethnicity was defined using self-reported data from primary care, supplemented with linked hospital admissions data where missing (recommended by Mathur et al. 2013 [16]). Patient were categorised into the major UK ethnicity groups: White, South Asian, Black, and Mixed/Other (combined due to low numbers). [17] For individuals with codes for multiple recorded ethnicities, we used the group with the highest number of recorded codes; if counts were equal we used the most recent code. Further details on this coding strategy can be found here: https://github.com/Exeter-Diabetes/CPRD-Codelists/blob/main/readme.md#ethnicity.

### Outcomes

The outcomes matched those of the original model development study. [5] The primary outcome comprised achieved 6-month HbA1c (closest measure to 6-months of treatment initiation, within a window of 3 to 15 months, on unchanged therapy). Secondary outcomes included 6-month weight change and treatment discontinuation within 6-months of initiation (as a proxy for non-tolerability). Each outcome was analysed using an outcome-specific cohort with complete cases for the outcome and all relevant baseline characteristics, as explained in sFigure 1.

### Model predictors and other clinical features

Extracted clinical features at drug initiation comprised: age, sex, diabetes duration, baseline HbA1c, BMI, eGFR, ALT, and the number of current, and ever, prescribed glucose-lowering drug classes.

### Statistical analysis

We implemented a 3-step approach for ethnicity-specific model optimisation and performance assessment.

#### Step 1: Determining the need for model updating in each ethnicity group and drug class

This assessment focused on evaluating the accuracy of predicted 6-month HbA1c outcomes from the original model [5] versus observed 6-month HbA1c outcomes and was executed separately for each ethnicity group and drug class. The requirement for model updating the original treatment selection model to improve prediction of 6-month HbA1c outcome by ethnicity was assessed using a closed testing procedure informed by Vergouwe et al. 2017 [18]. We employed a sequence of log-likelihood tests to determine whether the original treatment selection model required recalibration.

The original model (Model 1) was compared with two recalibrated models: recalibration-in-the-large (Model 2, updated model intercept) and intercept/slope recalibration (Model 3, updated intercept and overall calibration slope). For Model 2, updated intercepts for each drug class were calculated by fitting an intercept-only linear regression model, with the difference between observed HbA1c and predicted HbA1c from the original model as the outcome. The estimated intercept from this model represented the average difference between the observed and predicted HbA1c outcomes. Model 3 recalibration was performed by regressing the observed HbA1c outcome against the predicted HbA1c outcome from the original model

To test whether the updated ethnicity-specific recalibrated models led to improvements over the original model, three model comparisons were performed using a nominal p-value for α = 5%. [18] The first test compared the prediction performance of Model 1 (original model) versus the Model 2 (recalibration-in-the-large model). The Model 2 versus Model 1 test was performed using a log-likelihood test with m degrees of freedom, where m is the number of estimated parameters in the original model (intercept not considered). The second test compared Model 2 with Model 3 (updated intercept and slope), and the test was performed with m+1 degrees of freedom. [18]

We then applied the following decision rules to perform ethnicity-specific updates for predictions for each drug class:

- if the second test was significant, Model 3 (updated intercept and slope) was chosen,
- if the first but not the second test was significant, Model 2 (updated intercept) was chosen,
- if none of the tests were significant, Model 1 (original model) was chosen.

#### Step 2: Assessment of calibration accuracy of predicted differences by drug class in 6-month HbA1c

The calibration accuracy of the chosen ethnicity-specific treatment selection model for predicting differences between drug classes in 6-month HbA1c was analysed for each ethnicity subgroup by comparing the mean predicted HbA1c difference to the model-estimated observed HbA1c differences, across deciles of model-predicted HbA1c difference. As counterfactual outcomes for each therapy in individual patients were not available, the procedure for this model validation followed the original study. [5] Firstly, we predicted the achieved HbA1c on both treatments for each individual in the study cohort. Secondly, we calculated individual-level predicted HbA1c differences as the difference between SGLT2i and DPP4i model predictions for each patient. Thirdly, we stratified individuals within each ethnicity group based on deciles of predicted HbA1c difference. In the last step, we estimated average observed HbA1c differences between individuals receiving SGLT2i and DPP4i within decile group, with adjustments to account for the fact that the individuals receiving these two different drug classes were not matched. These observed differences were estimated from a linear regression model with achieved HbA1c as the outcome, received drug class as the main effect, and adjusted for baseline HbA1c, number of other current glucose-lowering drugs, number of glucose-lowering drug classes ever prescribed, month outcome HbA1c measurement was taken, eGFR, log of ALT, current age and baseline BMI (continuous features modelled as 3-knot restricted cubic splines).

#### Step 3: Estimation of unadjusted differences in 6-month HbA1c response (change from baseline in HbA1c), weight change and early discontinuation

Individuals in the study cohort were stratified into five subgroups defined using clinically relevant thresholds of predicted individual glycaemic benefit (benefit with SGLT2i versus DPP4i: ≥5 mmol/mol, 3-5 mmol/mol, 0-3 mmol/mol; benefit with DPP4i versus SGLT2i: 0-3 mmol/mol, 3-5 mmol/mol). For each subgroup, we estimated unadjusted mean glycaemic response, weight change, and discontinuation proportions by drug class.

All analyses were conducted using R (version 4.4.1), and all analysis code can be found online at: https://github.com/Exeter-Diabetes/CPRD-LauraJohn-SGLT2iDPPi-ethnicity. We followed TRIPOD prediction model reporting guidance. [19]

## Results

The study cohort included 145,556 drug initiations of SGLT2i (n = 57,749) and DPP4i (n = 87,807) (patient flowchart: sFigure 1). 78.5% (n = 114,287) were of White, 4.6% (n = 6,663) of Black, 14.4% (n = 20,969) of South Asian and 2.5% (n = 3,637) of Mixed/Other ethnicity. Baseline characteristics for the HbA1c outcome cohort are shown in Table 1.

**Table 1:** Baseline clinical characteristics of the study cohort for treatment selection model validation of HbA1c outcome. Baseline clinical characteristics for the secondary outcome cohorts are shown in sTable 1 and sTable 2. Data are mean (1 SD) for continuous variables.

|  | **DPP4i**<br>**(N = 61,440)** | **SGLT2i**<br>**(N = 36,047)** |
| --- | --- | --- |
| **Current age, years** | 62.8 (12.0) | 58.5 (10.4) |
| **Duration of diabetes, years** | 8.9 (6.7) | 9.2 (6.3) |
| **Sex** |  |  |
| Female | 23,652 (38.5%) | 13,578 (37.7%) |
| Male | 37,788 (61.5%) | 22,469 (62.3%) |
| **Ethnicity** |  |  |
| White | 48,832 (79.5%) | 28,497 (79.1%) |
| South Asian | 8,397 (13.7%) | 5,209 (14.5%) |
| Black | 2,780 (4.5%) | 1,409 (3.9%) |
| Mixed or other | 1,431 (2.3%) | 932 (2.6%) |
| **DPP4-inhibitor type** |  |  |
| Alogliptin | 12,923 (21.0%) | - |
| Linagliptin | 12,926 (21.0%) | - |
| Saxagliptin | 3,686 (6.0%) | - |
| Sitagliptin | 31,388 (51.1%) | - |
| Vildagliptin | 517 (0.8%) | - |
| **SGLT2-inhibitor type** |  |  |
| Canagliflozin | - | 6,489 (18.0%) |
| Dapagliflozin | - | 15,642 (43.4%) |
| Empagliflozin | - | 13,891 (38.5%) |
| Ertugliflozin | - | 25 (0.1%) |
| **Number of glucose-lowering drug classes ever prescribed** |  |  |
| 2 | 27,638 (45.0%) | 8,610 (23.9%) |
| 3 | 25,993 (42.3%) | 11,387 (31.6%) |
| 4+ | 7,809 (12.7%) | 16,050 (44.5%) |
| **Number of other current glucose-lowering drugs** |  |  |
| 0 | 3,473 (5.7%) | 1,182 (3.3%) |
| 1 | 36,017 (58.6%) | 14,578 (40.4%) |
| 2 | 21,228 (34.6%) | 16,286 (45.2%) |
| 3 | 722 (1.2%) | 4,001 (11.1%) |
| **Background therapy** |  |  |
| Metformin | 55,087 (89.7%) | 32,995 (91.5%) |
| Sulfonylurea | 22,267 (36.2%) | 12,959 (36.0%) |
| DPP4-inhibitor | - | 10,438 (29.0%) |
| SGLT2-inhibitor | 1,687 (2.7%) | - |
| Thiazolidinedione | 1,264 (2.1%) | 910 (2.5%) |
| GLP-1 receptor agonist | 201 (0.3%) | 1,847 (5.1%) |
| **Baseline biomarkers** |  |  |
| HbA1c mmol/mol | 72.2 (13.4) | 76.0 (14.3) |
| BMI, kg/m<sup>2</sup> | 31.8 (6.4) | 33.7 (6.7) |
| eGFR, mL/min per 1.73 m<sup>2</sup> | 88.6 (17.8) | 95.1 (14.5) |
| Alanine transaminase, IU/L | 3.3 (0.5) | 3.4 (0.5) |
| **HbA1c outcome** |  |  |
| HbA1c, mmol/mol | 63.6 (15.5) | 64.2 (14.1) |
| Month of HbA1c measure | 6.7 (2.8) | 6.6 (2.7) |

### A model update was required to adjust for a greater glycaemic response to DPP4i in people of non-White ethnicity

The model updating procedure identified greater observed average glycaemic response with DPP4i than predicted by the original model, which was most pronounced in individuals of non-White ethnicity. Model intercept recalibration (Model 2) was therefore performed to allow for greater DPP4i response for all ethnicity groups, as follows: White -1.6 mmol/mol; Black -3.0 mmol/mol; South Asian -2.6 mmol/mol; Mixed/Other -2.6 mmol/mol. For individuals initiating SGLT2i across non-White ethnicity groups, the original model fitted well, and no model updates were required. For the White ethnicity group initiating SGLT2i, a -0.9 mmol/mol intercept adjustment was applied. A summary of the full testing procedure results is provided in sTable 3.

### Model predicted and observed differences in HbA1c are accurate for all ethnicity groups

Figure 1a shows the distribution of the differential treatment effects predicted from the updated treatment selection model. In all ethnicity groups, a majority of individuals were predicted to have a higher glycaemic benefit on SGLT2i compared to DPP4i (63% to 79% across ethnicity groups). The average predicted response was 3.7 mmol/mol (95% CI 3.5 to 3.9) greater with SGLT2i than DPP4i for those of White ethnicity; this was greater than for those of South Asian (2.1 mmol/mol (95% CI 1.6 to 2.6)), Black (0.6 mmol/mol (95% CI 0.5 to 1.7)) and Mixed/Other (2.6 mmol/mol (95% CI 1.4 to 3.8)) ethnicity groups.

**Figure 1a:**
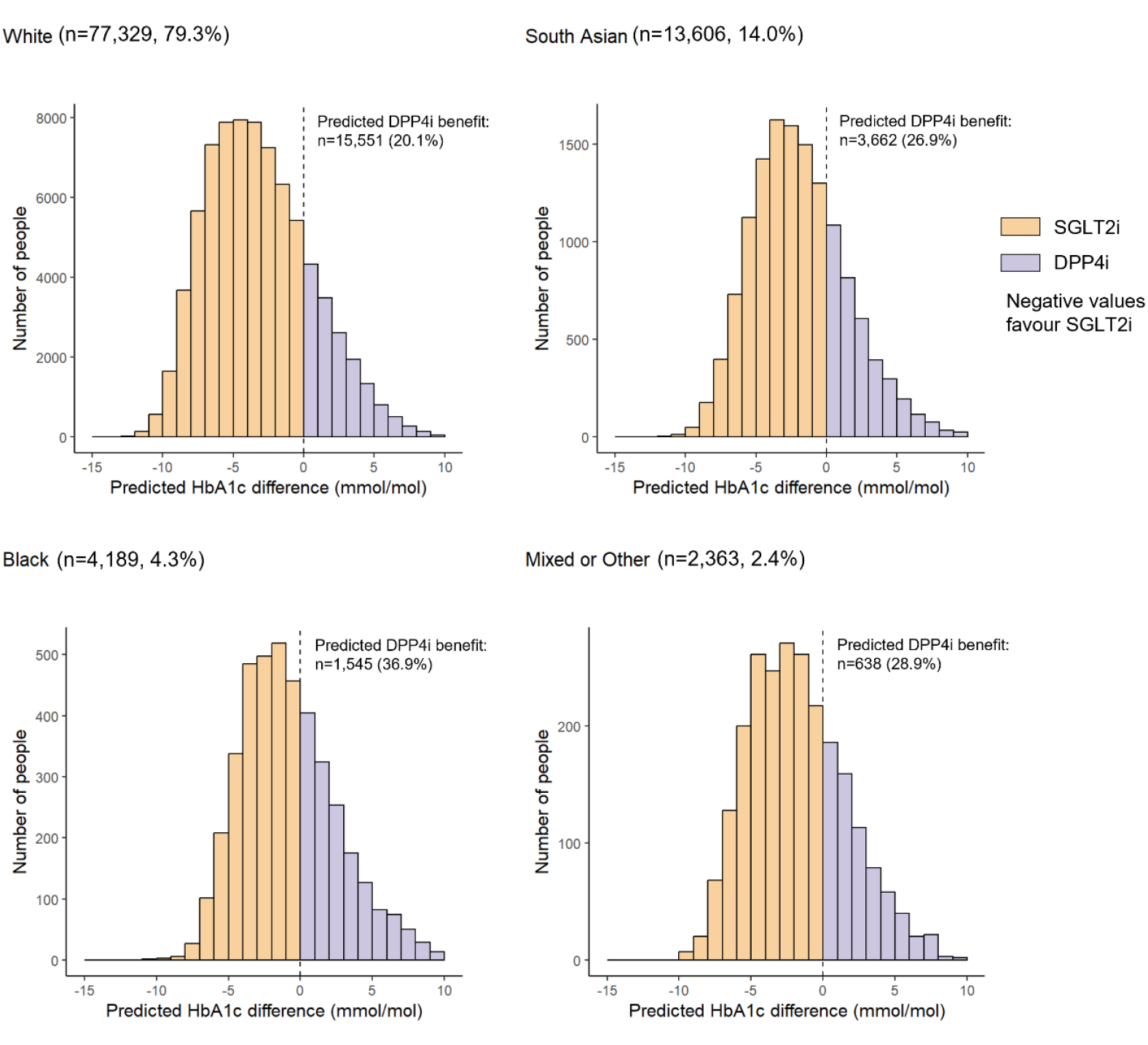
Distribution of predicted individual-level differential treatment effects for 6-month HbA1c of SGLT2i-inhibitor treatment compared to DPP4-inhibitor treatment, by UK ethnicity group. Negative values reflect a predicted 6-month HbA1c benefit on SGLT2-inhibitor treatment, positive values reflect a predicted 6-month HbA1c on DPP4-inhibitor treatment.

The results of the calibration assessment for the updated model are shown in Figure 1b. Within deciles of predicted individualised treatment effect, adjusted average HbA1c differences and predictions from the updated model were generally similar across all ethnicity groups, indicating that the predicted individualised treatment effects were well calibrated.

**Figure 1b:**
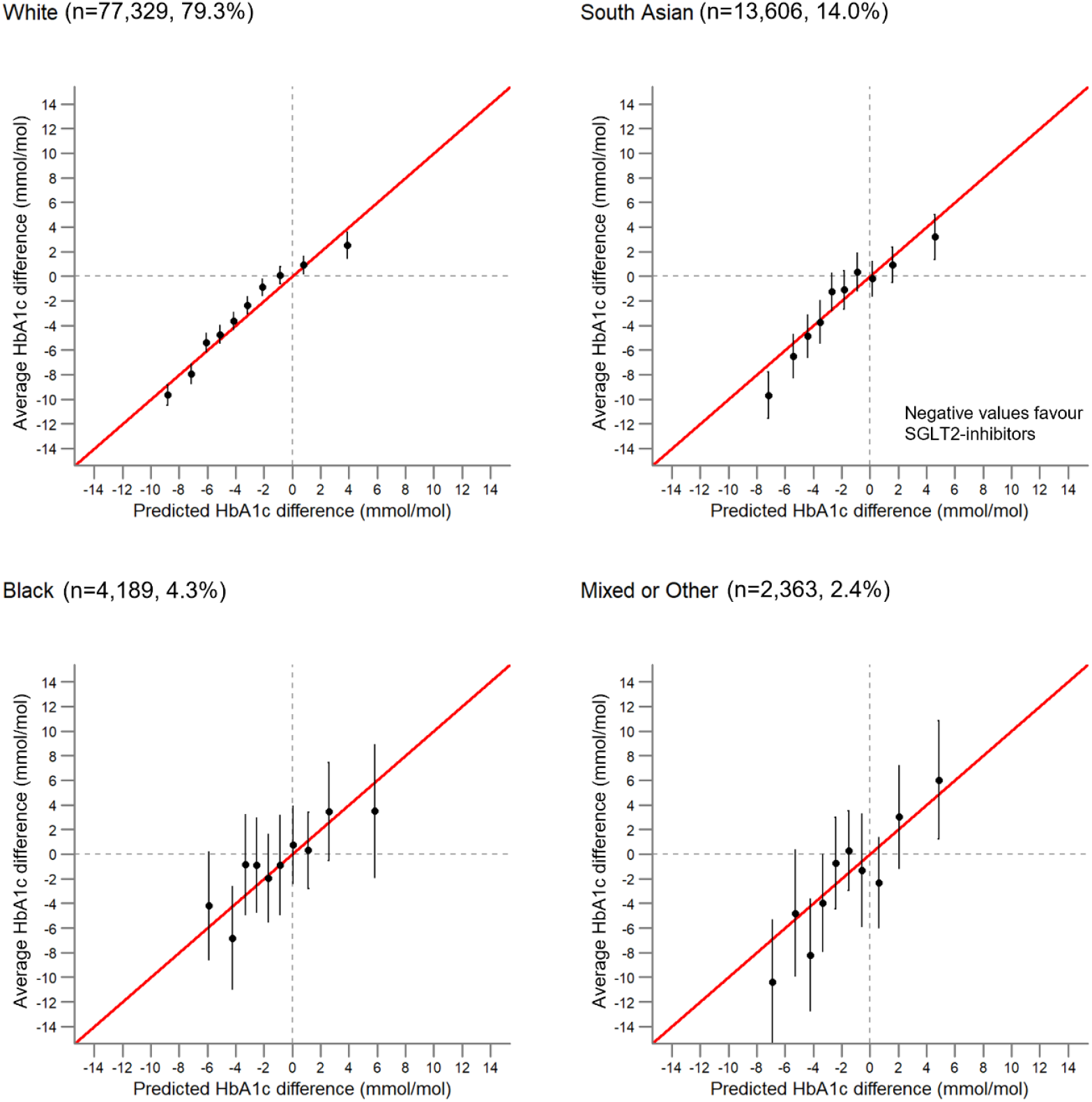
Calibration of predicted 6-month HbA1c benefit, by UK ethnicity group. Red lines represent perfect calibration. Point estimates represent treatment effects for subgroups defined by decile of predicted treatment benefit. Bars represent 95% confidence intervals. Negative values reflect a predicted 6-month HbA1c benefit on SGLT2-inhibitor treatment, positive values reflect a predicted 6-month HbA1c on DPP4-inhibitor treatment.^1^ ^1^ Average HbA1c differences are adjusted absolute mean differences in 6-month HbA1c outcome between individuals receiving each drug class.

### Across all ethnicity groups, the model identifies clinically meaningful differences in HbA1c response, including a subgroup with negligible response to SGLT2i

Figure 2a shows observed mean 6-month HbA1c response to each drug class in subgroups defined by predicted differential glycaemic response for each ethnicity group. Differences in predicted response between the two drug classes were broadly consistent with observed differences. Subgroups predicted to have a benefit on SGLT2i greater than 5 mmol/mol actually achieved a HbA1c benefit on SGLT2i of 6.2 mmol/mol (95% CI 5.8 to 10.4) higher compared to on DPP4i in the White ethnicity group, 3.1 mmol/mol (95% CI 1.0 to 12.9) in the Black ethnicity group, 6.3 mmol/mol (95% CI 5.1 to 12.9) in the South Asian ethnicity group, and 6.9 mmol/mol (95% CI 3.7 to 8.0) in the Mixed/Other ethnicity group. Individuals predicted to have a benefit on DPP4i of more than 3 mmol/mol achieved an average glycaemic benefit of 4.7 mmol/mol (95% CI 3.4 to 6.0) in the White ethnicity group, 4.2 mmol/mol (95% CI 0.4 to 8.0) in the Black ethnicity group, 5.0 mmol/mol (95% CI 3.2 to 6.8) in the South Asian ethnicity group, and 5.9 mmol/mol (95% CI 1.3 to 10.4) in the Mixed/Other ethnicity group higher than on SGLT2i.

**Figure 2a:**
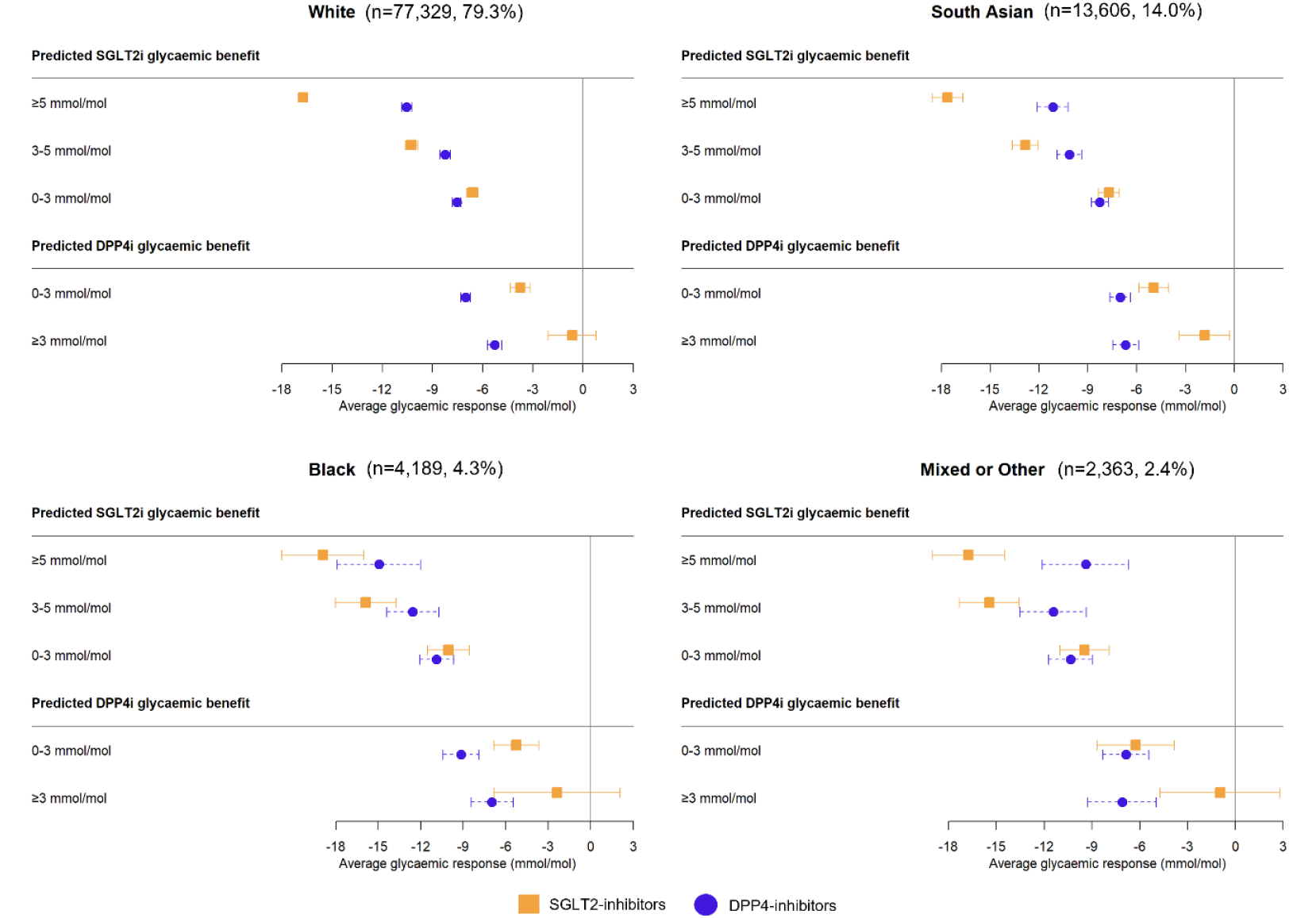
Observed 6-month HbA1c response in subgroups defined by predicted differential glycaemic response and UK ethnicity group. Point estimates represent unadjusted mean change from baseline in HbA1c. Bars represent 95% confidence intervals. sTable 4 reports underlying 6-month HbA1c response estimates.

Interestingly, in those predicted to have a DPP4i benefit of >3mmol/mol, this appeared to be driven by a lack of meaningful response to SGLT2is in this subgroup (-0.7 mmol/mol (95% CI -1.9 to 0.5) for White ethnicity, -3.4 mmol/mol (95% CI -7.0 to 0.2) for Black ethnicity, -1.5 mmol/mol (95% CI -3.0 to 0.0) for South Asian ethnicity and -0.8 mmol/mol (95% CI -4.4 to 2.8) for Mixed/Other ethnicity). This subgroup represented 6.6% of those of White, 8.5% of South Asians, 13.4% of Black, and 9.5% of Mixed/Other ethnicities (sTable 4).

### All ethnicity groups have a greater weight loss with SGLT2i versus DPP4i, but those with the greatest predicted glycaemic benefit with DPP4i have a higher rate of discontinuation if initiating SGLT2i

Figure 2b shows consistently higher weight reduction with SGLT2i versus DPP4i across subgroups defined by predicted differential glycaemic response for all ethnicity groups. Similarly, differences in patterns of early discontinuation were similar by ethnicity, with higher discontinuation for people initiating SGLT2i in those with a predicted benefit on DPP4i, although this was most pronounced for individuals of White ethnicity group (Figure 2c).

**Figure 2b:**
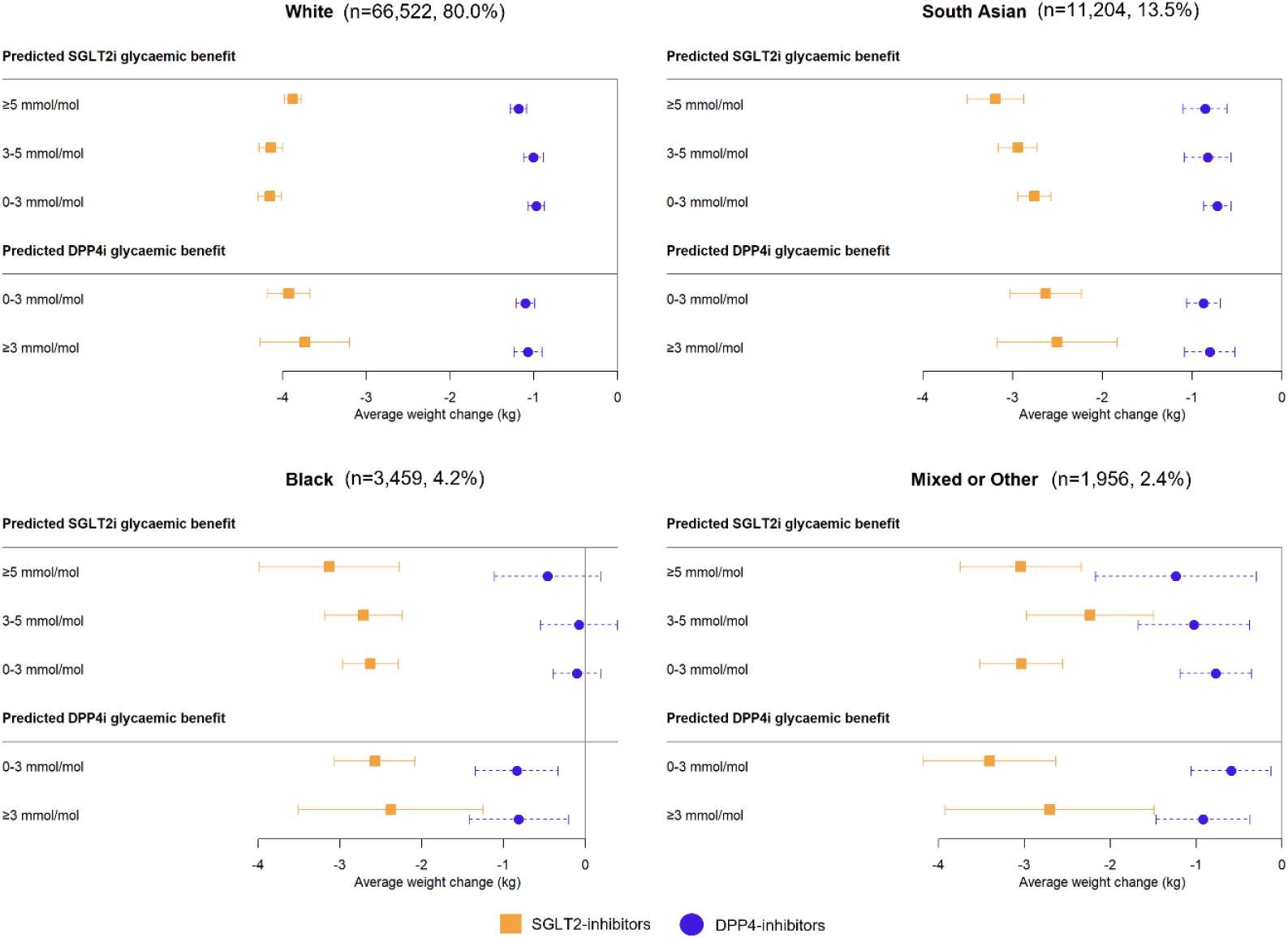
Observed 6-month weight change in subgroups defined by predicted differential glycaemic response and UK ethnicity group. Point estimates represent unadjusted mean change from baseline in weight, including all individuals with valid baseline data for the treatment selection model and a valid weight outcome. Bars represent 95% confidence intervals. sTable 5 reports underlying 6-month weight change estimates.

**Figure 2c:**
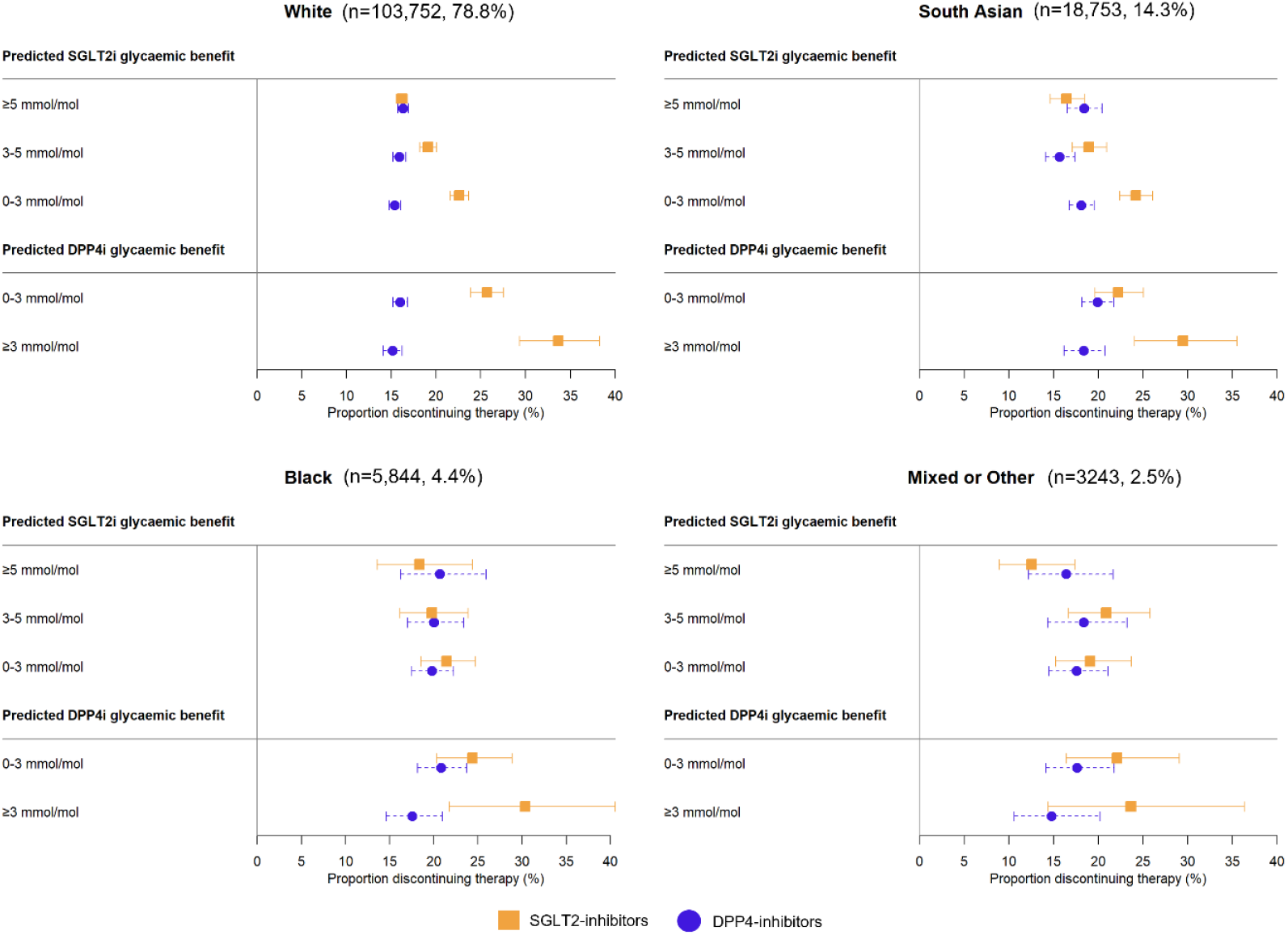
Proportion of individuals discontinuing treatment within 6-months in subgroups defined by predicted differential glycaemic response and UK ethnicity group. Point estimates are unadjusted, including all individuals with valid baseline data for the treatment selection model and 3 additional months of follow up to confirm treatment was truly discontinued. Bars represent 95% confidence intervals. sTable 6 reports underlying 6-month discontinuation estimates.

## Discussion

In this external validation study, we show that a model designed to determine the optimal choice of SGLT2i and DPP4i therapies to maximise glycaemic response in people with type 2 diabetes is accurate across major UK ethnicity groups. Modest recalibration was required to reflect a greater average glycaemic response with DPP4i than predicted by the original model, of around 2-3 mmol/mol for South Asian, Black, and Mixed/Other ethnicity groups. No model updates were required for SGLT2i initiations across all non-White ethnicity groups. Importantly, good performance was achieved without adjustment of the effect of individual model predictors themselves, suggesting that clinical factors altering the response to both therapies are consistent across ethnicity groups. The application of the updated model identified an interesting patient subgroup, representing 6-13% of individuals across the ethnicity groups, with a significant HbA1c response to DPP4i but only a very limited response to SGLT2i inhibitors. Weight change and short-term discontinuation patterns were consistent across ethnicities, with greater weight loss associated with SGLT2i and higher rates of discontinuation in individuals initiating SGLT2i who were predicted to derive a greater glycaemic benefit from DPP4i. Overall, findings support the potential clinical deployment of the SGLT2i-DPP4i model for targeting treatment in diverse populations. Our methodology also provides an example of a principled approach for the external validation of treatment selection model to support translational precision medicine.

There is limited evidence from previous studies about differential glycaemic reductions with SGLT2i versus DPP4i across ethnicity groups. Previous trial meta-analysis report DPP4i to have a greater glycaemic efficacy in people of Asian ethnicity than in other ethnicity groups [14, 20], as we found in this study. Potential mechanisms for these findings include a mediation of DPP4i efficacy by clinical factors associated with insulin sensitivity, such as differences between ethnicity groups in BMI, β-cell function (HOMA-%B), or the amount of visceral fat. A further suggestion is that there may be differences in the pharmacokinetic properties of DPP4i in Asian and non-Asian groups due to variations in body size. [14, 20] Despite this enhanced DPP4i response, our results show that SGLT2i still have a greater glycaemic benefit compared to DPP4i for most people across all ethnicity groups. Similar to previous studies, our results did not show clear treatment effect heterogeneity with SGLT2i by ethnicity group. [9-12, 21] Overall, the DPP4i specific ethnicity differences identified in this study, although relatively modest on average, highlight the importance of future research to identify differential T2D drug effects by ethnicity and to interrogate potential underlying mechanisms.

Considering the secondary outcomes we assessed, SGLT2i have been consistently shown to lead to greater weight loss compared with DPP4i across different populations. [22, 23] This finding was consistent in our study across all ethnicity groups, including when stratified by glycaemic benefit. Whilst limited previous studies have shown lower rates of discontinuation with SGLT2i versus DPP4i [24, 25], they did not analyse differential discontinuation risk by ethnicity.

A major strength of our study is the use of a large and diverse population-based cohort, which included near complete data on self-reported ethnicity, and incorporation of a substantial number of individuals from non-White ethnic groups allowing for well-powered subgroup analysis even for differential treatment effects. External validation of the original model [5] was possible as the study population from the database CPRD Aurum was collected from different practices using the EMIS clinical system compared to with CPRD Gold data which was used to develop the treatment selection model. [15] Our study population also relied on a robust open-source pipeline for processing and analysing electronic health record data, supporting research reproducibility. [26] Recent analyses by the UK Office for National Statistics have highlighted ecosystem-wide challenges in the accuracy of routinely collected ethnicity data [27], and thus, it is important to recognise that completeness alone is an imperfect metric for quality in this context. No linkage to the 2022 ONS census ethnicity data currently exists for CPRD (a potential limitation of this study, as this would be considered the gold standard record for self-reported ethnicity). For when these linkages become available, we have provided a robust, fully-documented, open-source pipeline for processing and analysing electronic health record data, to support reproducibility [26], and replication of our results. Furthermore, our study explores weight change and early treatment discontinuation as key short-term outcomes beyond HbA1c, for which evidence of treatment effect heterogeneity is lacking. Another strength of this study is the application of a testing procedure informed by Vergouwe et al. 2017 [18] to assess the prediction performance of models based on a sequence of log-likelihood tests. This procedure makes it possible to rationally adapt prediction models for treatment selection to study populations that differ from the population in which the model was initially developed. Importantly, the procedure allowed us to flexibly update the model, allowing multiple potential updating strategies to be fairly evaluated. A limitation of our study is that it was not possible to validate individual differential treatment effects due to a lack of counterfactual outcome measurements. [28] We therefore did not establish causal effects between exposure and outcome of interest but focused on evaluating associations. This observational study also did not address unmeasured confounding which leaves the possibility for residual unmeasured confounding bias, although reassuringly the model performed well when previously validated in multinational clinical trials. [5] A further limitation is that we did not evaluate heterogeneity in cardiorenal outcomes, which was also not evaluated in the previous study, although our methods could be extended to these longer-term outcomes in future work to allow a more holistic consideration of treatments that may best benefit individual patients. A final limitation is our focus on validation of the two-drug SGLT2i and DPP4i treatment model, considering we recently published a five-drug class model also including GLP-1 receptor agonists, sulfonylureas, and thiazolidinediones. [29] The current study provides the methodological framework to perform future validation of the five-drug class model both by ethnicity in the UK when sufficient additional longitudinal data are available, and in other populations and settings beyond the UK.

Results of this study suggest that individualised treatment selection models relying on simple routine clinical features have clinical utility for targeting treatment choice in diverse populations. Hence, this study highlights the clear potential utility of treatment selection models as equitable low-cost approaches which can improve T2D care worldwide. Our novel framework for model testing and validation enables robust evaluation of treatment selection models in a new population, a crucial step in ensuring treatment selection algorithms are fit for informing clinical decisions. Accordingly, extending the validation framework developed in this study to evaluate our recently proposed five drug class treatment selection model [29] in diverse populations is a key area for future research.

## Conclusion

A SGLT2i-DPP4i treatment selection model accurately predicts differences in HbA1c responses in all major UK ethnicity groups. Model evaluation shows that in all ethnicity groups, a majority of individuals were predicted to have a greater glycaemic reduction on SGLT2i compared with DPP4i. The benefit of SGLT2i is however less marked in non-white ethnicity groups due to a greater response to DPP4i. Individualised treatment selection models have great potential to provide a low-cost, pragmatic approach to T2D precision medicine with applicability across diverse populations.

## Data Availability

CPRD Autum data are available by application to the CPRD Independent Scientific Advisory Committee. R code to preproduce the analysis in this paper is available at https://github.com/Exeter-Diabetes/CPRD-LauraJohn-SGLT2iDPPi-ethnicity.

## Declarations

## Acknowledgements

This article is based in part on data from the Clinical Practice Research Datalink obtained under licence from the UK Medicines and Healthcare products Regulatory Agency. CPRD data is provided by patients and collected by the NHS as part of their care and support. Approval for CPRD data access and the study protocol was granted by the CPRD Independent Scientific Advisory Committee (eRAP protocol number: 24_004747).

## Author contribution

LMG JMD, BS and AGJ designed the study and developed the analysis strategy. LMG and JMD conducted the analysis. PC supported the application of the testing procedure and KGY prepared the CPRD data and constructed the T2D cohort. LMG drafted the original version of the paper which all authors helped to edit. BAM, RRH and NS provided critical revisions of the manuscript. ERP and ATH provided valuable clinical insights and helped interpret the results. All authors participated in editing and revising the manuscript draft and have read and approved the final version. LMG and JMD are the guarantors of this work and as such, had full access to all the data in the study and take responsibility for the integrity of the data and the accuracy of the data analysis.

