## supplementary material for "Validation of a treatment selection algorithm for optimal choice of SGLT2 and DPP4 inhibitor therapies in people with type 2 diabetes across major UK ethnicity groups"

Laura M Güdemann (PhD)[1], Katherine G Young (PhD)[1], Pedro Cardoso (PhD)[1], Bilal A Mateen (MBBS) (Prof)[2], Rury R Holman (F.Med.Sci) (Prof)[3,4], Naveed Sattar (MD) (Prof)[5], Ewan R Pearson (PhD) (Prof)[6], Andrew T Hattersley (DM) (Prof)[1], Angus G Jones (PhD) (Prof)[1], Beverley M Shields (PhD)[1], John M Dennis (PhD)[1], on behalf of the MASTERMIND consortium

**Institutions:**

[1] Clinical and Biomedical Sciences, University of Exeter Medical School, Exeter, UK

[2] School of Life Sciences, University of Birmingham, Birmingham, UK

[3] Diabetes Trials Unit, Radcliffe Department of Medicine, University of Oxford, Oxford, UK

[4] NIHR Oxford Biomedical Research Centre, Churchill Hospital, Oxford, UK

[5] Institute of Cardiovascular and Medical Sciences, University of Glasgow, Glasgow, UK

[6] Division of Diabetes, Endocrinology and Reproductive Medicine, Ninewells Hospital and Medical School, University of Dundee, Dundee, UK

**Corresponding author**

John M. Dennis

Address: Clinical and Biomedical Sciences, University of Exeter Medical School, Exeter, United Kingdom. Tel: +44 7734 940921

### Supplementary material

#### sFigure 1: Flowchart

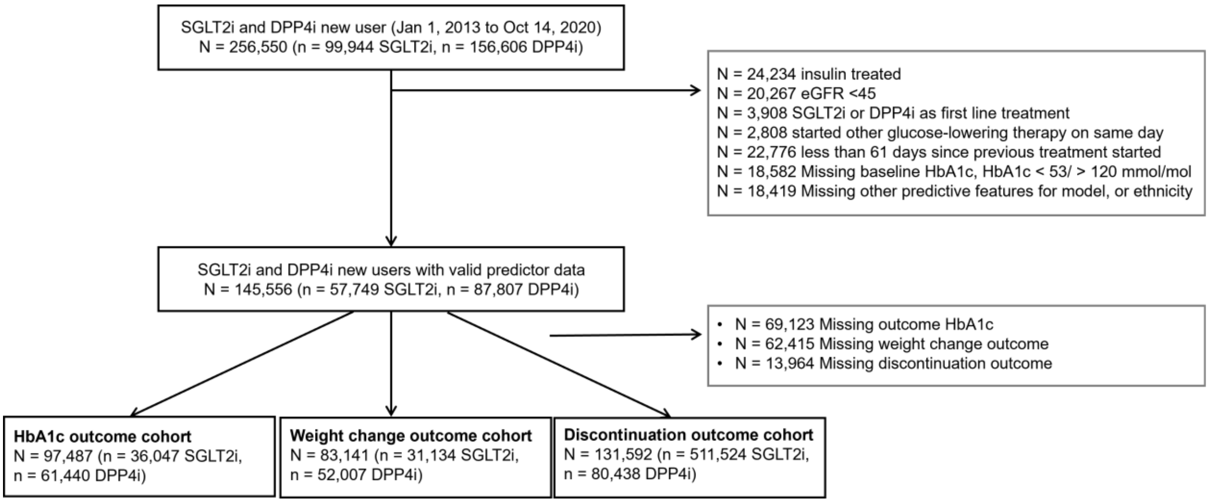

**sTable 1: Baseline clinical characteristics of the weight change outcome specific study cohort.** Data are mean (1 SD) for continuous variables.

| | **DPP4i (N = 52,007)** | **SGLT2i (N = 83,141)** |
|---|---|---|
| **Current age, years** | 62.7 (11.9) | 58.4 (10.3) |
| **Duration of diabetes, years** | 8.8 (6.6) | 9.1 (6.3) |
| **Sex** | | |
| Female | 23,652 (38.5%) | 11,677 (37.5%) |
| Male | 32,041 (61.6%) | 19,457 (62.5%) |
| **Ethnicity** | | |
| White | 41,676 (80.1%) | 24,846 (79.8%) |
| South Asian | 6,878 (13.2%) | 4,326 (13.9%) |
| Black | 2,267 (4.4%) | 1,192 (3.8%) |
| Mixed or other | 1,186 (2.3%) | 770 (2.5%) |
| **DPP4-inhibitor type** | | |
| Alogliptin | 10,960 (21.1%) | - |
| Linagliptin | 10,811 (20.8%) | - |
| Saxagliptin | 3,231 (6.2%) | - |
| Sitagliptin | 26,554 (51.1%) | - |
| Vildagliptin | 451 (0.9%) | - |
| **SGLT2-inhibitor type** | | |
| Canagliflozin | - | 5,644 (18.1%) |
| Dapagliflozin | - | 13,632 (43.8%) |
| Empagliflozin | - | 11,837 (38.0%) |
| Ertugliflozin | - | 21 (0.1%) |
| **Number of glucose-lowering drug classes ever prescribed** | | |
| 2 | 23,482 (45.2%) | 7,422 (23.8%) |
| 3 | 21,980 (42.3%) | 9,791 (31.4%) |
| 4+ | 6,545 (12.6%) | 13,921 (44.7%) |
| **Number of other current glucose-lowering drugs** | | |
| 0 | 2,772 (5.3%) | 994 (3.2%) |
| 1 | 30,594 (58.8%) | 12,606 (40.5%) |
| 2 | 18,046 (34.7%) | 14,078 (45.2%) |
| 3 | 595 (1.1%) | 3,456 (11.1%) |
| **Background therapy** | | |
| Metformin | 46,870 (90.1%) | 28,576 (91.8%) |
| Sulfonylurea | 18,838 (36.2%) | 11,123 (35.7%) |
| DPP4-inhibitor | - | 9,005 (28.9%) |
| SGLT2-inhibitor | 1,409 (2.7%) | - |
| Thiazolidinedione | 1,075 (2.1%) | 776 (2.5%) |
| GLP-1 receptor agonist | 167 (0.3%) | 1,644 (5.3%) |
| **Baseline biomarkers** | | |
| HbA1c mmol/mol | 71.9 (13.3) | 75.8 (14.3) |
| BMI, kg/m² | 31.9 (6.4) | 33.8 (6.7) |
| eGFR, mL/min per 1.73 m² | 88.7 (17.7) | 95.2 (14.5) |

| | | |
|---|---|---|
| Alanine transaminase, IU/L | 32.3 (19.3) | 34.9 (20.1) |
| **Weight outcome** | | |
| Achieved weight (kg) | 90.4 (20.4) | 93.8 (20.9) |

**sTable 2: Baseline clinical characteristics of the discontinuation outcome specific study cohort.** Data are mean (1 SD) for continuous variables.

| | **DPP4i (N = 41122)** | **SGLT2i (N = 24077)** |
|---|---|---|
| **Current age, years** | 62.6 (12.1) | 58.5 (10.5) |
| **Duration of diabetes, years** | 8.91 (6.69) | 9.29 (6.30) |
| **Sex** | | |
| Female | 31617 (39.3%) | 19845 (38.8%) |
| Male | 48821 (60.7%) | 31309 (61.2%) |
| **Ethnicity** | | |
| White | 63498 (78.9%) | 40254 (78.7%) |
| South Asian | 11242 (14.0%) | 7511 (14.7%) |
| Black | 3775 (4.7%) | 2069 (4.0%) |
| Mixed or other | 1923 (2.4%) | 1320 (2.6%) |
| **DPP4-inhibitor type** | | |
| Alogliptin | 16922 (21.0%) | - |
| Linagliptin | 17042 (21.2%) | - |
| Saxagliptin | 4757 (5.9%) | - |
| Sitagliptin | 41054 (51.0%) | - |
| Vildagliptin | 663 (0.8%) | - |
| **SGLT2-inhibitor type** | | |
| Canagliflozin | - | 9244 (18.1%) |
| Dapagliflozin | - | 22084 (43.2%) |
| Empagliflozin | - | 19767 (38.6%) |
| Ertugliflozin | - | 59 (0.1%) |
| **Number of glucose-lowering drug classes ever prescribed** | | |
| 2 | 35809 (44.5%) | 11968 (23.4%) |
| 3 | 34040 (42.3%) | 16014 (31.3%) |
| 4+ | 10589 (13.2%) | 23172 (45.3%) |
| **Number of other current glucose-lowering drugs** | | |
| 0 | 5092 (6.3%) | 1991 (3.9%) |
| 1 | 47043 (58.5%) | 20289 (39.7%) |
| 2 | 27191 (33.8%) | 22286 (43.6%) |
| 3 | 1112 (1.4%) | 6588 (12.9%) |
| **Background therapy** | | |
| Metformin | 71273 (88.6%) | 46134 (90.2%) |
| Sulfonylurea | 28812 (35.8%) | 18824 (36.8%) |
| DPP4-inhibitor | - | 15492 (30.3%) |
| SGLT2-inhibitor | 2341 (2.9%) | - |
| Thiazolidinedione | 1761 (2.2%) | 1441 (2.8%) |
| GLP-1 receptor agonist | 380 (0.5%) | 2766 (5.4%) |

| | | |
|---|---|---|
| **Baseline biomarkers** | | |
| HbA1c mmol/mol | 72.6 (13.8) | 76.2 (14.6) |
| BMI, kg/m² | 31.9 (6.51) | 33.6 (6.76) |
| eGFR, mL/min per 1.3 m² | 88.8 (17.9) | 95.0 (14.8) |
| Alanine transaminase, IU/L | 32.3 (19.3) | 34.9 (20.1) |
| **Discontinuation outcome** | | |
| Discontinuation, yes | 13356 (16.6%) | 10026 (19.6%) |

**sTable 3: Results of the model updating testing procedure.**

| Cohort | | N | P-values | | Final model chosen | log-likelihood | Updated intercept (95% CI)* |
|---|---|---|---|---|---|---|---|
| Treatment | Ethnicity | | Test 1 | Test 2 | | | |
| DPP4-inhibitors | White | 48,832 | < 0.0001 | 0.14 | Updated intercept | -194,208 | -1.6 (-1.7, -1.5) |
| SGLT2-inhibitors | White | 28,497 | < 0.0001 | 0.38 | Updated intercept | -110,538 | -0.9 (-1.0, -0.7) |
| DPP4-inhibitors | Black | 2,780 | < 0.0001 | 0.79 | Updated intercept | -11,644 | -3.0 (-3.6, -2.4) |
| SGLT2-inhibitors | Black | 1,409 | < 0.0001 | 1 | Original | -5,765 | NA |
| DPP4-inhibitors | South Asian | 8,397 | 1 | 1 | Updated intercept | -33,076 | -2.6 (-2.8, -2.3) |
| SGLT2-inhibitors | South Asian | 5,209 | 1 | 1 | Original | -20,360 | NA |
| DPP4-inhibitors | Mixed/Other | 1,431 | < 0.0001 | 1 | Updated intercept | -5,733 | -2.6 (-3.3, -2.0) |
| SGLT2-inhibitors | Mixed/Other | 932 | 1 | 1 | Original | -3,625 | NA |

Test 1 compared the prediction performance of Model 1 (original model) versus Model 2 (recalibration-in-the-large model). Test 2 compared Model 2 with Model 3 (updated intercept and slope). If the second test was significant, Model 3 was chosen, if the first but not the second test was significant, Model 2 was chosen, and if none of the tests were significant, Model 1 was chosen.
*A negative value represents greater response than predicted by the original model
